# Tiny dLIF: A Dendritic Spiking Neural Network Enabling a Time-Domain Energy-Efficient Seizure Detection System

**DOI:** 10.1101/2024.05.23.24307841

**Authors:** Luis Fernando Herbozo Contreras, Leping Yu, Zhaojing Huang, Ziyao Zhang, Armin Nikpour, Omid Kavehei

## Abstract

Epilepsy poses a significant global health challenge, driving the need for reliable diagnostic tools like scalp electroencephalogram (EEG), subscalp EEG, and intracranial EEG (iEEG) for accurate seizure detection, localization, and modulation for treating seizures. However, these techniques often rely on feature extraction techniques such as Short Time Fourier Transform (STFT) for efficiency in seizure detection. Drawing inspiration from brain architecture, we investigate biologically plausible algorithms, specifically emphasizing time-domain inputs with low computational overhead. Our novel approach features two hidden layer dendrites with Leaky Integrate-and-Fire (dLIF) spiking neurons, containing fewer than 300K parameters and occupying a mere 1.5 MB of memory. Our proposed network is tested and successfully generalized on four datasets from the USA and Europe, recorded with different front-end electronics. USA datasets are scalp EEG in adults and children, and European datasets are iEEG in adults. All datasets are from patients living with epilepsy. Our model exhibits robust performance across different datasets through rigorous training and validation. We achieved AUROC scores of 81.0% and 91.0% in two datasets. Additionally, we obtained AUPRC and F1 Score metrics of 91.9% and 88.9% for one dataset, respectively. We also conducted out-of-sample generalization by training on adult patient data, and testing on children’s data, achieving an AUROC of 75.1% for epilepsy detection. This highlights its effectiveness across continental datasets with diverse brain modalities, regardless of montage or age specificity. It underscores the importance of embracing system heterogeneity to enhance efficiency, thus eliminating the need for computationally expensive feature engineering techniques like Fast Fourier Transform (FFT) and STFT.

## 1 Introduction

Epilepsy is a neurological condition characterized by recurrent seizures, impacting millions of individuals globally. Approximately 30% of cases exhibit resistance to conventional Anti-Epileptic Drugs (AEDs), resulting in drug-resistant epilepsy. Despite extensive efforts in developing and testing AEDs, there has been limited advancement in enhancing their efficacy [1]. The unpredictable nature of seizures poses significant risks to patients’ quality of life, employment status, and overall well-being, including potential hazards such as falls and sudden unexpected death in epilepsy [2, 3]. Implementing a dependable system for precise seizure detection and quantification could substantially improve decision-making processes, treatment strategies, and disease management, ultimately improving patient outcomes. Technologies for electronically sensing the brain include electroencephalogram (EEG) and electrocorticography (ECoG) aka. iEEG (intracranial EEG), capturing the brain’s electrical activity from the scalp or head surface, often non-invasively or using electrodes sitting directly on the brain’s surface (subdurally/epidurally), respectively. iEEG obviously provides superior spatial and temporal resolution as well as Signal-to-Noise Ratio (SNR) but accessing the brain surface has been a major challenge surgically [4]. iEEG, if it is done for relatively long-term use, has several applications, including brain-machine interface [5], but it is often used as a presurgical method to locate the seizure focal points in patients with epilepsy [6]. ECoG or iEEG can also be recorded as endovascular, which is a breakthrough in the delivery mechanism of electrodes to the brain, albeit often without a chance for the device’s explantation [7, 8, 9, 10]. The use of Artificial Intelligence (AI) in brain signal analysis is shown to be reasonably successful for efficient seizure detection [11, 12, 13, 14, 15]. AI applications in EEG monitoring are limited by hospital resources and hardware/software constraints. Current AI focus in EEG monitoring with fewer electrodes, automated channel reduction, and suggests multi-modal data fusion [16]. These AI methods encompass diverse approaches, including traditional and embedded AI. Traditional AI usually operates on robust Graphics Processing Units (GPU) clusters, facilitating ultra-fast computations and EEG data analysis, while embedded AI involves integrating AI capabilities directly in or close to edge sensors, enabling more real-time analysis and decision-making on the device itself. This approach is especially beneficial for continuous monitoring and early seizure detection, as it minimizes data transmission requirements, making it well-suited for long-term monitoring scenarios with on-device learning.

### 1.1 Background

Researchers are actively developing AI models for seizure detection. For instance, studies have combined Independent Component Analysis (ICA) and Short Time Fourier Transform (STFT) for pre-processing, followed by Convolutional-Long Short Term Memory (ConvLSTM) blocks, achieving an Area Under the Receiver Operating Characteristic Curve (AUROC or AUC) of 0.84 in the largest epilepsy dataset in United States [17]. Similarly, by utilizing domain adaptation and STFT pre-processing, authors has reported an AUC of 0.75 ± 0.03 [18]. Moreover, a study has introduced anchored-STFT to enhance temporal-spectral resolution trade-off in EEG analysis [19]. Despite their strong performance, these models often demand significant memory and power resources, making them impractical for deployment on edge devices. Neuromorphic AI, inspired by the brain, differs significantly from the traditional architecture based on von Neumann, which utilizes separated memory and processing units, consuming power during data transmission. Instead, a neuromorphic chip employs physical neurons interconnected by physical synapses, implementing collocated memory processing in a non-volatile manner. This design drastically reduces the necessity to move data across the circuit, leading to substantial improvements in speed and energy efficiency [20]. While the biophysical model has dendrites, axons and receptors, the current models only consider the somas. Dendrites are crucial for neuronal computation due to their ability to act as semi-independent thresholding units, generating local dendritic Spikes (dSpikes). These spikes are produced by voltage-gated mechanisms and influence synaptic input integration and plasticity [21]. Dendritic mechanisms operate across multiple timescales, enabling complex computations such as coincidence detection, filtering, input segregation, nonlinear processing, and logical operations [22, 23]. Therefore, dendrites are essential for accurately modeling neuronal integration and output at the single-cell level, contributing significantly to the computational power of neural networks. However, the current theoretical framework for modelling dendrite properties consists of complex equations with numerous free parameters, making it mathematically intractable and impractical for use in Spiking Neural Networks (SNNs). Simulators have been proposed that enable dendrites to operate semi-independently from the soma and perform complex functions, enhancing the computational capabilities of the model [24], but not being scaled to SNNs.

### 1.2 The proposed system

In this study, we proposed tiny dendritic Leaky Integrate-and-Fire (dLIF) SNN that embraces dendritic computations by representing them as small RC circuits to provide heterogeneity to the network [25]. This integration contributes to improved computational efficiency and balanced performance. The model is based on two hidden layers, with dendritic inputs that act as rich inputs, considering how biological neurons receive, process, and transmit information. Notably, the simplicity of this model allows for memory utilization of approximately 1.5 MB with less than 300K parameters.

### 1.3 Novelty and significance

This work leverages the potentials of dLIF models for seizure detection, which enables:

i. Direct analysis in the time domain without relying on traditional time-frequency transformation techniques like STFT or FFT. This approach simplifies the pre-processing analysis, potentially reducing power and hardware requirements.
ii. Tiny model by its minimal requirement of only two hidden layers, enhancing its efficiency below 300K parameters.
iii. Down-sampling scalp-EEG signal (i.e 250 Hz to 125 Hz) yield similar performance, paving the way for rapid, energy-efficient training systems.
iv. Efficacy across different brain activity recordings is demonstrated through rigorous testing using both scalp-EEG and iEEG datasets.
v. Out-of-sample seizure detection by training with adult patients montages and testing in children montages.

## 2 Methods

### 2.1 Datasets

There are 4 datasets used in this work: the Temple University Hospital (TUH) Corpus scalp-EEG, the Children’s Hospital Boston (CHB-MIT) dataset, the Freiburg Hospital (FB), and the intracranial-EEG EPILEPSIAE dataset. Fig. 2 illustrates a summary of the TUH dataset used, providing key statistics related to its data. This dataset is divided into two categories: one set of training and one set of validation, offering a comprehensive overview of the dataset’s contents. It includes information such as the number of patients with and without seizure sessions. It also shows the inter-ictal and ictal periods used (Background and Seizure duration). With its large volume of files, the TUH dataset from America offers vast data and served as a meaningful scalp-EEG dataset as it is the largest dataset worldwide. Most of these patients are also adults. We performed out-of-sample generalization in the CHB-MIT dataset for a non-montage and age-dependent systems. We randomly selected 17 patients; the most significant channels were the same number as the channels trained in the TUH dataset. We compared with previous studies that utilize different model structures, with time-frequency domain and in-sample testing. We explored further how our model can recognize and train with iEEG signals. Thereby, we utilize the FB and EPILEPSIAE datasets. We trained and tested in-sample in 14 patients as these datasets do not possess the same montages and different amounts of channels across the EPILEPSIAE, varying from 30 to 120 electrodes, utilizing a sampling rate of 256 Hz. The EPILEPSIAE dataset contains high-quality, long-term EEG, intracranial EEG, and concurrently recorded ECG data. Intracranially implanted strips, grids, and/or stereotactically (stereo-EEG) implanted depth electrodes are used in patients with invasive recordings. The EPILEPSIAE dataset possessed very long interictal periods (background data between ictal events), which is a much more realistic iEEG dataset and makes our false negatives highly reliable. The FB dataset contains 21 intractable epilepsy patients with iEEG recordings. In this dataset, data was recorded from 6 selected electrodes, at a sampling rate of 256 Hz, where three are epileptogenic regions, and the rest are from other remote areas. 12 patients with 311.4 hours recording length are tested because of the availability of the dataset. A summary detailed of these datasets can be seen in Table 1.

**Table 1.**
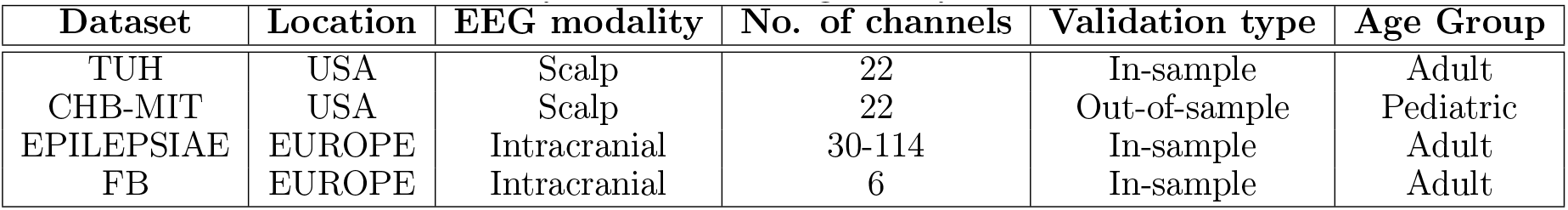
Summary of datasets being employed in this work.

| Dataset | Location | EEG modality | No. of channels | Validation type | Age Group |
| --- | --- | --- | --- | --- | --- |
| TUH | USA | Scalp | 22 | In-sample | Adult |
| CHB-MIT | USA | Scalp | 22 | Out-of-sample | Pediatric |
| EPILEPSIAE | EUROPE | Intracranial | 30-114 | In-sample | Adult |
| FB | EUROPE | Intracranial | 6 | In-sample | Adult |

**Figure 1.**
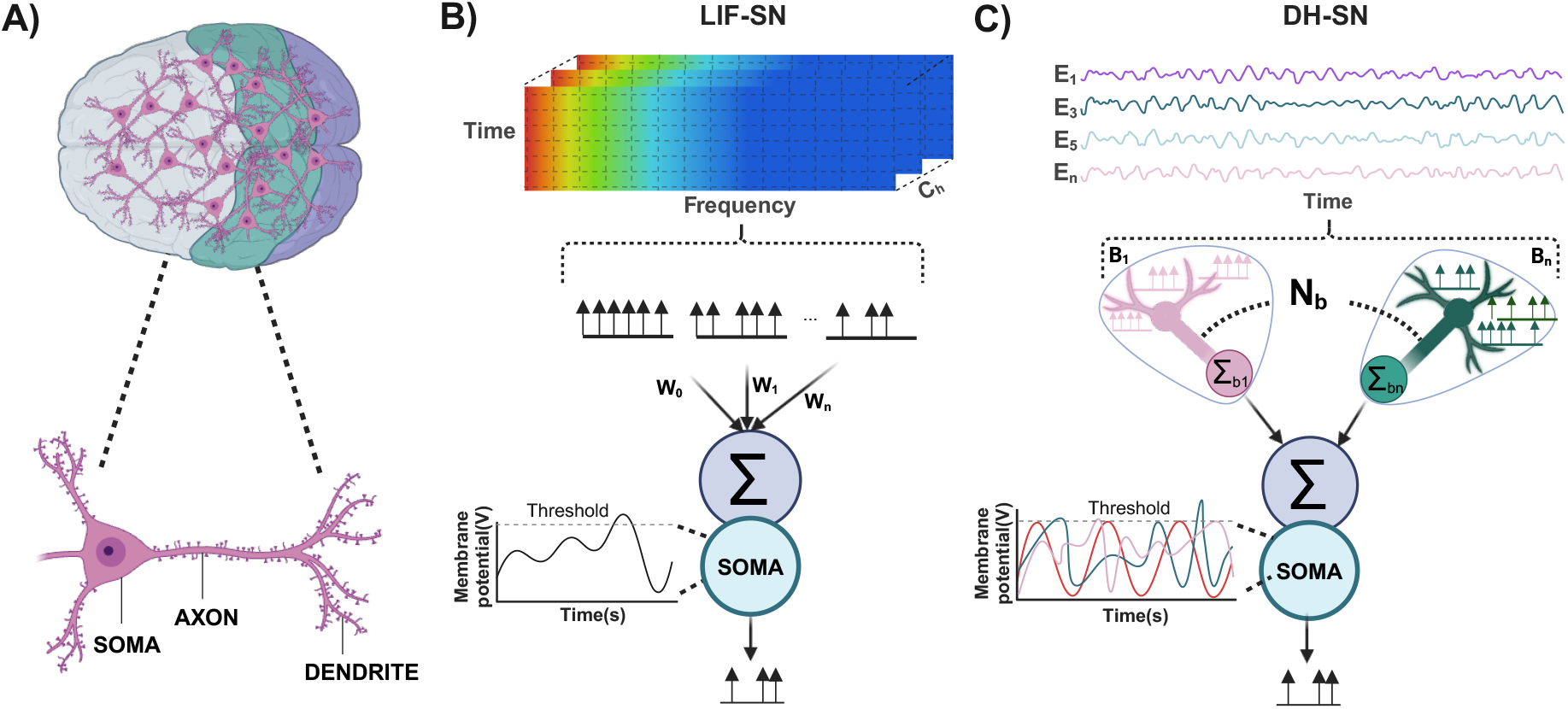
Biological Inspired Neuron Model. The main components of the brain are neurons, which have different parts as the dendrites, soma, and axon (A). Different types of biological-inspired neural networks, like LIF Neurons, are built from the Soma and do not consider the dendrites as part of their system. Therefore, it minimizes the heterogeneity of an actual brain. Those models often use Time-Frequency Analysis to enhance efficiency (B). However, this analysis can be computational and power costly; therefore, adding heterogeneity by incorporating dendrites in the LIF-Spiking Neurons allows time-domain series processing. B_1-n_: Branches, N_b_: Number of Branches, W_n_ = Synaptic Weights.

**Figure 2.**
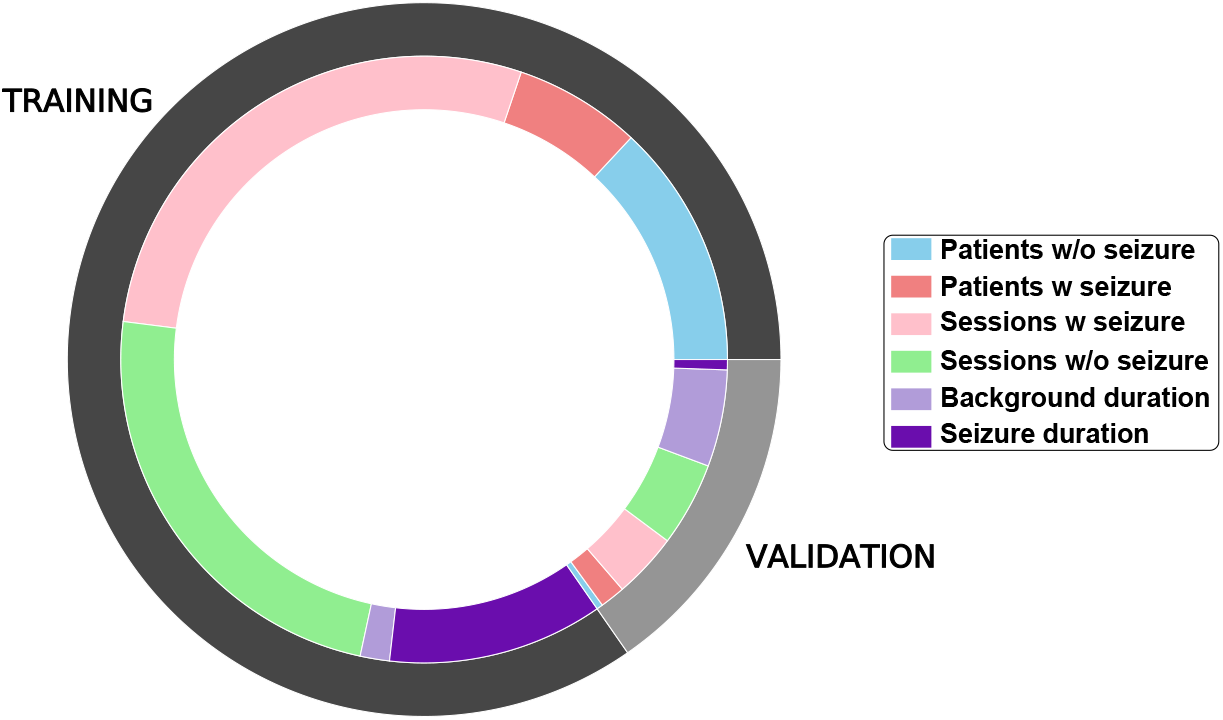
An overview of the TUH dataset used for training and validation.

### 2.2 Pre-Processing

We utilized the Independent Component Analysis (ICA) technique to address the challenges related to scalp-EEG data artifacts removal. Initially, the EEG signals were split into 12-second segments, and the ICA algorithm was applied to decompose the signals into 19 independent components using Blind Source Separation (BSS). ICA separates EEG signals into statistically independent components, represented in Equ. (1),

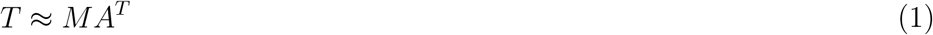

where *T* contains the EEG data, *M* contains the time information, and *A* contains the weights for topographic maps. We used Pearson correlation to identify independent sources strongly associated with eye movement, which was detected from two EEG channels, ‘FP1’ and ‘FP2’. These identified sources related to eye movement were removed from the independent components, resulting in EEG signals free from eye movement artifacts. Subsequently, we implemented power-line noise removal for both the FB and iEEG-EPILEPSIAE datasets at 50 Hz, employing a notch filter sourced from the MNE Package. For the TUH and CHB-MIT datasets, we applied a notch filter at 60 Hz. This measure was imperative due to detecting noise within the specified frequency range via power spectrum analysis.

### 2.3 SNN structure

#### 2.3.1 dLIF Spiking Neuron Structure

The dLIF neuron model enhances the classic LIF-based spiking neuron by introducing multi-timescale memory on dendrites, governed by Equ. (2), (3), (4) where u represents the soma’s membrane potential, *β* is the timing factor, *R* is membrane resistance, *d* is the dendritic branch index, and *u*_th_ is the firing threshold [25].

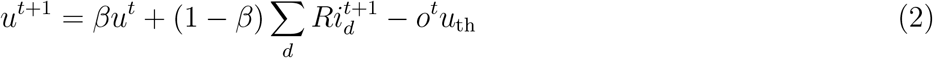

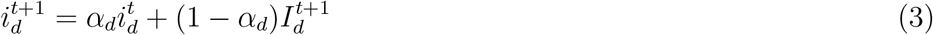

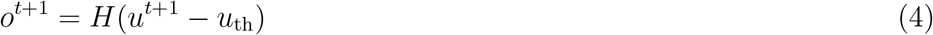

The Heaviside function *H*(*Heaviside function H(*) regulates spiking. Synaptic input on the *d*_th_ dendritic branch is the sum of feedforward and recurrent inputs as shown in Equ. (5), defined by sparse vectors *W*_*d*_ and *U*_*d*_ [25].

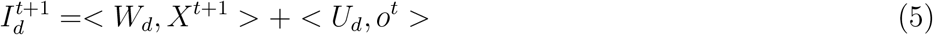

Extending this to dLIF-SNN involves incorporating layer information as shown in Equs. (6), (7), (8), with synaptic currents on the d_*th*_ branch as shown in Equ. (9), determined by matrix forms of feed-forward and recurrent synaptic weights (*W*_*d*_ and *U*_*d*_), which are again sparse, considering only valid synapses connected to dendritic branches in the next layer [25].

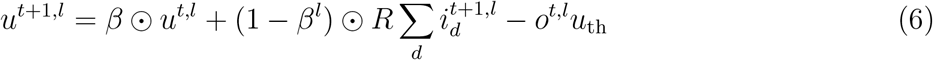

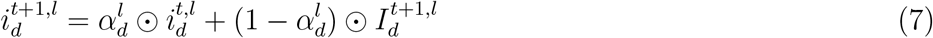

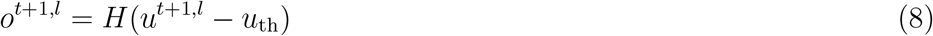

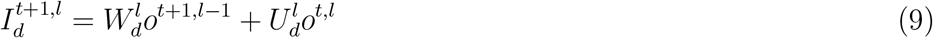

#### 2.3.2 Learning rule

The Backward Propagation Through Time (BPTT) algorithm has been adopted better to handle the unique characteristics of spiking neural networks. During training, model parameters such as synaptic weights (*W*) and timing factors 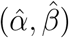 are automatically learned to optimize network performance based on the loss function [25]. The Equ. (10), (11), (12) shows the details of the BPTT where *δ* denotes the gradient of the loss function L concerning specific variables [25]. The BPTT for dLIF-SNN utilizes gradient descent with the chain rule to update parameters while dealing with non-differentiable spiking activities using a soft multi-Gaussian curve shown in Equ. (13) as a surrogate gradient function, with parameters like *γ, h* which will influence the magnitude, *±, s* which will influence the width [25].

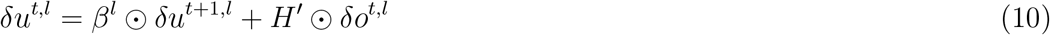

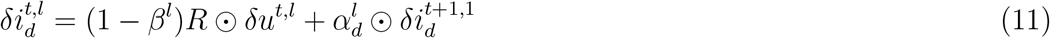

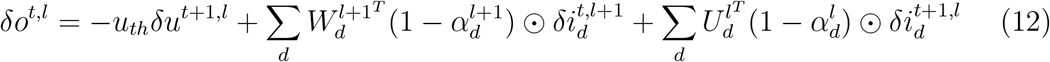

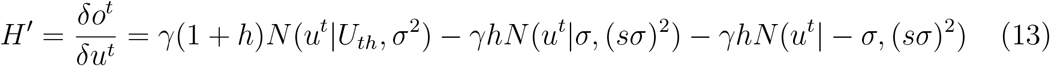

Gradients of parameters are shown in Equ. (14), (15), (16) are computed using this surrogate gradient function, particularly around the firing threshold (u_*th*_) where neurons emit spikes, enabling efficient training and improved performance of dLIF-SNNs [25].

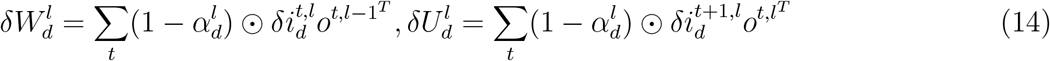

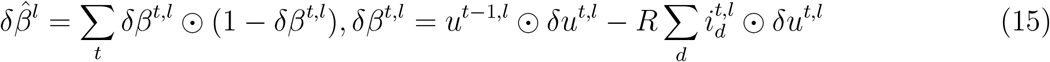

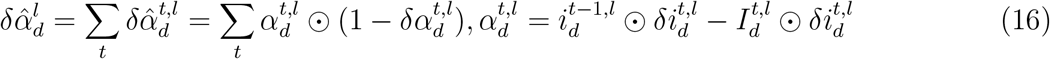

### 2.4 Implementation details and Performance metrics

We employed cross-entropy as our loss function and a softmax function of the model’s predictions to assess different performance metrics. We utilized the area under the receiver operating characteristic curve (AUC-ROC or AUROC). Specifically, The AUC captures both the sensitivity and specificity in a threshold-free score. We also provide precision, recall, and F1-score metrics to evaluate the model performance across different datasets. Precision measures the proportion of correctly identified positive cases among all cases predicted as positive by the model. Recall measures the proportion of correctly identified positive cases among all actual positive cases in the dataset. The F1-score is the harmonic mean of precision and recall. It provides a single metric that balances both precision and recall. In our experiments, it is worth noting that we performed in-sample testing with the TUH, FB, and iEEG-EPILEPSIAE datasets. However, we performed out-of-sample testing in the CHB-MIT dataset. A detailed diagram of the system is represented in Fig. 3. The model was trained and tested on a V100 GPU.

**Figure 3.**
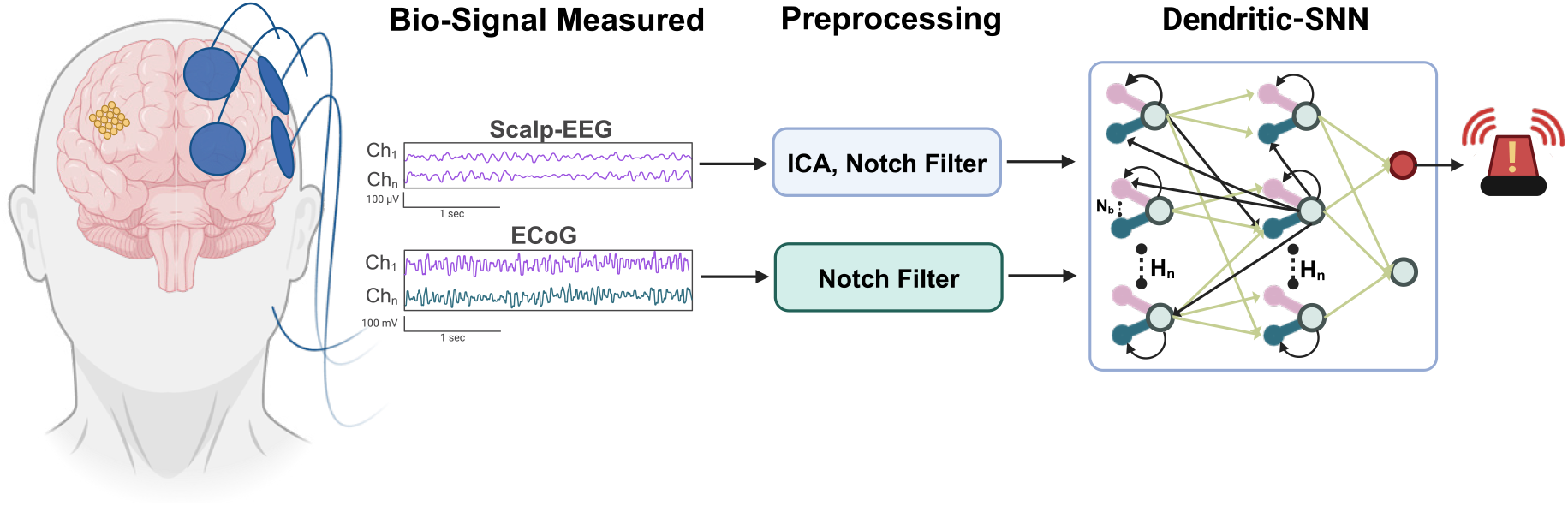
Neuromorphic Detecting System using dSNN. Bio-signals such as scalp-EEG and iEEG are preprocessed using a notch filter for power-line noise removal or ICA to separate blind-source components and then fed into a single two-layer dLIF for detecting seizures. N_b_: Number of branches (4), H_n_: Hidden Neurons (100).

## 3 Experimental Results

### 3.1 Scalp-EEG

#### 3.1.1 Training and Validation

Following the methodology outlined earlier, our model was trained and validated using the TUH dataset. We successfully attained an impressive AUC score of 81.4%. Our findings, as illustrated in Fig. 4 (A), highlight the superior performance of our spiking model compared to non-spiking models. Notably, our model’s performance rivals existing approaches within the time and time-frequency domains, with fewer layers and memory needs, and in a spiking fashion. Table 2 provides a comprehensive overview of the model performance across a range of metrics.

**Table 2.** TUH Results for down-sampling test. It is worth noting that a decrease of 50% in the original sampling frequency leads to similar performance.

| EEG Sampling Rate | AUROC | AUPRC | Precision | Recall | F1-Score |
| --- | --- | --- | --- | --- | --- |
| 250 Hz | 81.4 | 90.1 | 84.4 | 81.3 | 83.1 |
| 125 Hz | 81.0 | 89.2 | 84.2 | 74.9 | 79.2 |

**Figure 4.**
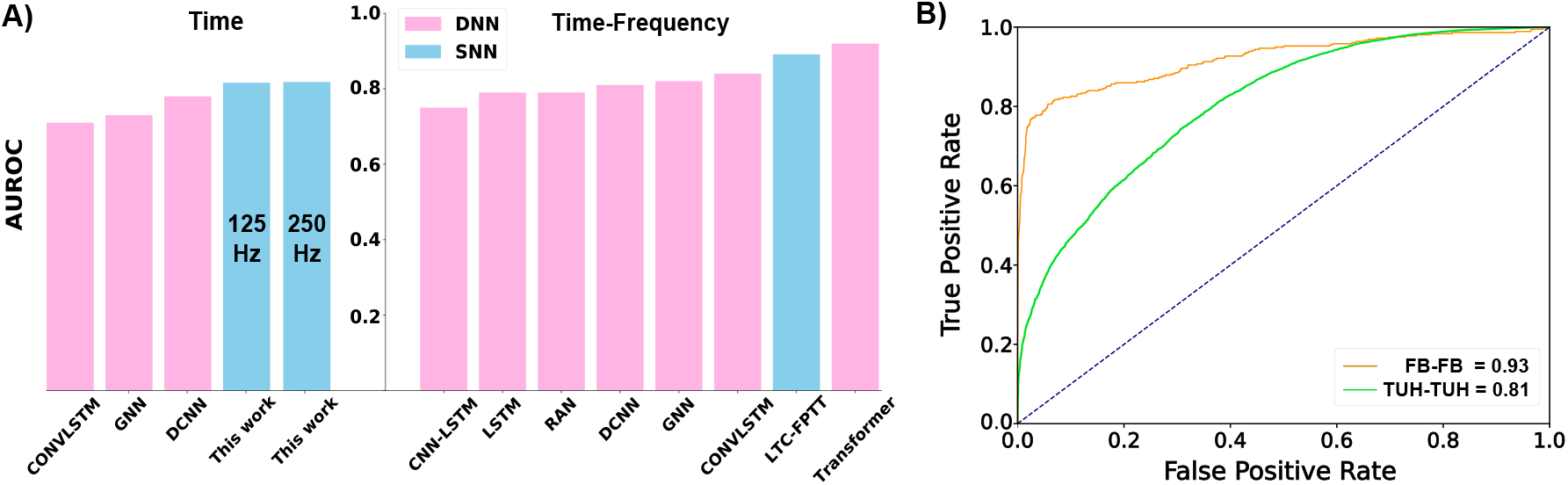
Model Performance Results. We assessed our model’s effectiveness against others using TUH Dataset, with a 12 s window Size and with Time or Time-Frequency Inputs (A). AUC-ROC in-sample testing results for both TUH and FB datasets shows an optimal balance between True Positive Rate and False Positive Rate (B).

#### 3.1.2 Preliminary findings in down-sampling the signal leads to similar performances

In our investigation, we examined the effects of reducing the sampling rate in the TUH dataset on our model’s performance. Impressively, our results showcased a remarkable level of robustness, with minimal drops in performance observed across key metrics such as AUROC, AUPRC, and Precision. Notably, while there was a slight decline of approximately 7% in Recall and 4% in F1-Score, the overall performance remained commendable. These findings support the hypothesis that our model prioritizes detection biomarkers located in lower frequencies over higher frequencies. This assertion is further substantiated by previous research [26]. Our results are represented in Table 2.

#### 3.1.3 Out-of-Sample generalization in children: Non-Montage and Non-Age Specific

This study investigated out-of-sample generalization in children, employing a non-montage and non-age-specific approach. Our research methodology involved rigorous testing utilizing the CHB-MIT dataset, a widely recognized repository for pediatric EEG data. Unlike previous models predominantly operating in the time-frequency domain, our novel approach focuses solely on the time domain, offering a unique perspective on EEG analysis in pediatric populations. Notably, our model adopts a spiking paradigm, further distinguishing it from existing methodologies. Importantly, we emphasize the significance of out-of-sample validation, diverging from the conventional in-sample evaluations commonly observed. Through meticulous analysis, we reveal how our framework, emphasizing time-domain evaluation and in-sample testing, differs from existing methodologies. Importantly, while our model does not demonstrate superiority in all aspects, it offers valuable insights by considering spiking models and temporal input data often overlooked in prior research. Results can be observed in Table 3, which demonstrate the need for embracing biological neural networks.

**Table 3.** CHB-MIT Results. Comparison of AUC-ROC across different models with time-frequency *vs* time-domain input (ours) and in sample *vs* out-sample (ours).

|  | In-Sample |  |  | Out-of-Sample |
| --- | --- | --- | --- | --- |
|  | Time-Frequency Domain |  |  | Time Domain |
| Patient ID | CNN [27] | Conv-LSTM [27] | SNN-Conv-LSTM [27] | dLIF-SNN |
| 1 | 100.0 | 100.0 | 96.4 | 71.9 |
| 3 | 100.0 | 100.0 | 92.3 | 51.1 |
| 5 | 100.0 | 100.0 | 95.3 | 66.4 |
| 6 | 94.9 | 93.4 | 88.8 | 51.1 |
| 7 | 97.0 | 96.5 | 92.0 | 87.3 |
| 8 | 99.4 | 98.2 | 72.5 | 79.8 |
| 9 | 99.7 | 100.0 | 96.1 | 98.3 |
| 10 | 100.0 | 100.0 | 97.5 | 86.6 |
| 11 | 98.8 | 98.9 | 81.2 | 98.6 |
| 15 | 99.9 | 99.7 | 73.0 | 50.7 |
| 17 | 88.2 | 91.0 | 87.7 | 88.1 |
| 18 | 98.5 | 96.9 | 86.0 | 63.0 |
| 19 | 95.4 | 99.8 | 95.9 | 72.0 |
| 20 | 97.1 | 97.7 | 87.9 | 68.8 |
| 21 | 99.3 | 100.0 | 92.5 | 65.7 |
| 22 | 100.0 | 100.0 | 96.6 | 95.6 |
| 23 | 100.0 | 100.0 | 95.9 | 95.0 |
| **Average** | 98.13 | 98.36 | 89.9 | 75.9 |

### 3.2 Intracranial-EEG

#### 3.2.1 Training and Validation

We conducted extensive analyses using data from both the FB and EPILEPSIAE datasets. Our examination involved multiple patients from the FB dataset, where we assessed our model’s performance in the time-frequency domain and compared it with alternative models. Surprisingly, even when utilizing only temporal input, our model exhibited efficacy on par with non-SNN and conventional SNN methods, achieving an impressive average AUROC score of 93.4. For the EPILEPSIAE dataset, we performed multiple training as each patient exhibited different channel numbers. Our study yielded notable outcomes across various evaluation metrics, including AUPRC, Sensitivity (Recall), Precision, and F1 Score, with impressive values of 92.0, 85.0, 95.0, and 89.0, respectively. These results, along with detailed comparative findings presented in Table 4, Fig. 4(B) and in Table 5, underscore the promising capabilities of our proposed approach in the analysis of iEEG signals.

**Table 4.** FB Results. Comparison of AUC-ROC across different models with time-frequency vs time-domain(ours).

|  | Time-Frequency Domain |  |  | Time Domain |
| --- | --- | --- | --- | --- |
| Patient ID | CNN [27] | Conv-LSTM [27] | SNN-Conv-LSTM [27] | dLIF |
| 1 | 100.0 | 100.0 | 100.0 | 100.0 |
| 3 | 100.0 | 100.0 | 93.7 | 80.2 |
| 4 | 100.0 | 100.0 | 99.9 | 96.1 |
| 5 | 85.0 | 84.8 | 84.4 | 100.0 |
| 6 | 86.8 | 90.6 | 92.4 | 53.3 |
| 14 | 80.5 | 92.6 | 85.6 | 100.0 |
| 15 | 93.3 | 99.1 | 89.5 | 100.0 |
| 16 | 93.4 | 96.1 | 85.7 | 99.0 |
| 17 | 100.0 | 100.0 | 97.3 | 94.9 |
| 18 | 100.0 | 100.0 | 98.5 | 99.1 |
| 20 | 94.8 | 99.1 | 94.5 | 99.2 |
| 21 | 98.8 | 97.5 | 91.4 | 99.7 |
| **Average** | 94.4 | 96.7 | 92.7 | 93.4 |

**Table 5.** iEEG-EPILEPSIAE Results. Training and testing occur independently for each patient.

| Patient ID | Gender | NoE | AUPRC | Sensitivity | Precision | F1 Score |
| --- | --- | --- | --- | --- | --- | --- |
| 1 | F | 66 | 78.1 | 71.2 | 70.0 | 71.1 |
| 2 | F | 30 | 100.0 | 100.0 | 100.0 | 100.0 |
| 3 | M | 114 | 96.1 | 92.3 | 100.0 | 96.1 |
| 4 | F | 75 | 97.2 | 93.1 | 100.0 | 96.2 |
| 5 | M | 82 | 80.3 | 50.3 | 100.0 | 67.1 |
| 7 | M | 60 | 98.1 | 92.1 | 100.0 | 96.2 |
| 8 | M | 38 | 60.2 | 50.1 | 68.0 | 58.1 |
| 9 | M | 92 | 99.2 | 97.2 | 100.0 | 98.1 |
| 10 | M | 56 | 94.1 | 75.3 | 100.0 | 86.6 |
| 11 | F | 46 | 100.0 | 100.0 | 100.0 | 100.0 |
| 12 | F | 124 | 99.1 | 95.1 | 100.0 | 97.1 |
| 13 | F | 62 | 98.0 | 88.3 | 100.0 | 94.2 |
| 14 | F | 121 | 95.4 | 89.2 | 97.3 | 93.1 |
| 15 | M | 58 | 91.1 | 93.1 | 89.1 | 91.2 |
| **Average** | - | - | 91.9 | 84.8 | 94.6 | 88.9 |

M: Male, F: Female, NoE: Number of invasive electrodes.

## 4 Discussion

Our next step is to incorporate this biologically plausible algorithm for a seizure prediction system and enhance its robustness for continuous learning and low computational preprocessing [28]. We foresee that this algorithm will be robust against catastrophic forgetting as it does not depend on back-propagation, and this phenomenon is present in back-propagation domains [29]. BPTT updates RNN parameters on an instance by back-propagating the error in time over the entire sequence length, leading to poor trainability due to the well-known gradient explosion/decay phenomena. Given the exceptional performance of Forward Propagation Through Time (FPTT) in previous studies with EEG data for seizure detection, we will integrate this approach into our model training framework [30] to deal with the challenges of BPTT. Although this training system takes considerable computation time per epoch, it is still more efficient than proposed delay hardware dendritic systems [31].

## 5 Conclusion

In this study, we assessed the advantages of compartmental models that enabled the analysis of effective time-domain. By only utilizing basic pre-processing and non-power consuming techniques such as power-line noise in real-world scenarios of iEEG datasets such as FB and EPILEPSIAE, and ICA on scalp-EEG datasets as the meaningful TUH dataset, we have demonstrated the capabilities to exhibit efficient performance while maintaining a low-power consumption spectrum. By only utilizing two hidden dendritic layers, we believe this study shows the possibility of more embedded AI applications for long-term data, demonstrating that more dynamic and biological neural networks are necessary. Future studies will merge liquid-time constant spiking neurons with dendritic neurons for more dynamic features.

## Data Availability

The TUH dataset is publicly available https://isip.piconepress.com/projects/tuh_eeg/html/downloads.shtml. The EPILEPSIAE dataset is available at cost via this http://www.epilepsiae.eu/project_outputs/european_database_on_epilepsy. The Children's Hospital Boston dataset is publicly available https://physionet.org/content/chbmit/1.0.0. The FB data used in this study used to be available openly via this https://epilepsy.uni-freiburg.de/freiburg-seizure-prediction-project/eeg-database, but it seems it no longer accepts registration.

https://isip.piconepress.com/projects/tuh_eeg/html/downloads.shtml

http://www.epilepsiae.eu/project_outputs/european_database_on_epilepsy

https://physionet.org/content/chbmit/1.0.0

https://epilepsy.uni-freiburg.de/freiburg-seizure-prediction-project/eeg-database

## 6 Acknowledgement

Luis Fernando Herbozo Contreras would like to acknowledge the partial support of the Faculty of Engineering Research Scholarship provided by The University of Sydney. Zhaojing Huang would like to acknowledge the support of the Research Training Program (RTP) provided by the Australian Government. Omid Kavehei acknowledges the support provided by The University of Sydney through a SOAR Fellowship and Microsoft’s support through a Microsoft AI for Accessibility grant.

## 7 Code Access

If you have any questions regarding access to the code employed in this research paper, please direct your inquiries to the corresponding author.

## 8 Data Accessibility

The TUH dataset is publicly available here. The EPILEPSIAE dataset is available at cost via this link. The Children’s Hospital Boston dataset is publicly available here. The FB data used in this study used to be available openly via this link, but it seems it no longer accepts registration.

## 9 Conflict of Interest Statement

The authors affirm that they have no conflicts of interest to disclose, including financial and non-financial considerations.

## Notes

### Competing Interest Statement

The authors have declared no competing interest.

